# Dietary diversity moderates household economic inequalities in the double burden of malnutrition in Tanzania

**DOI:** 10.1101/2023.10.10.23296857

**Authors:** Sanmei Chen, Yoko Shimpuku, Takanori Honda, Dorkasi L. Mwakawanga, Beatrice Mwilike

## Abstract

**Background:** Improved food availability and a growing economy in Tanzania may insufficiently decrease preexisting nutritional deficiencies and simultaneously increase overweight within the same individual, household, or population, causing a double burden of malnutrition (DBM). We investigated economic inequalities in DBM at the household level, expressed as a stunted child with an overweight/obese mother, and the moderating role of dietary diversity in these inequalities.

**Methods:** We used cross-sectional data of 2,867 children (aged 6–23 months) and their mothers (aged 15–49 years) from the 2015–2016 Tanzania Demographic and Health Survey. The participants were categorized into two groups based on their dietary diversity score: achieving and not achieving minimum dietary diversity. We assessed the interaction effect between household wealth and dietary diversity on DBM and examined the association between household wealth and DBM in subgroups based on achieving minimum dietary diversity. Logistic regression models with sample weights and restricted cubic spline functions were used for the analysis.

**Results:** The prevalence of DBM was 5.6% (SD=0.6) and significantly varied by regions (ranging from 0.6%–12.2%). Significant interaction was observed between dietary diversity and household wealth index (*p for* interaction = 0.01). The prevalence of DBM monotonically increased with greater household wealth among those who did not achieve minimum dietary diversity (*p* for trend = 0.002; however, but this association was attenuated in those who achieved minimum dietary diversity (*p* for trend = 0.15), particularly for the richest households (*p* = 0.43). Similar results were observed when modeling the household wealth index score as a continuous variable (odds ratio [95% confidence interval]: 2.05 [1.32-3.19] for non-achievers of minimum dietary diversity, 1.39 (0.76─2.54) for achievers).

**Conclusions:** Greater household wealth was associated with higher odds of DBM in Tanzania; however, achieving minimum dietary diversity may mitigate the negative impact of household economic status on DBM.

**What is already known on this topic?:** The double burden of undernutrition and obesity is increasing in low- and middle-income countries and has serious and lasting developmental and socioeconomic impact.

Household economic inequalities have been linked to the double burden of malnutrition (DBM) at the household level. However, existing evidence is conflicting and limited in Tanzania.

Diversified diet may mitigate the potential adverse impact of household economic inequalities on DBM; however, evidence is scarce.

**What this study adds?:** This study is one of the few attempts to explore economic inequalities in DBM at the household level in Tanzania by considering the moderating role of dietary diversity in these inequalities.

The prevalence of DBM varies regionally and is unequally distributed across levels of household wealth nationwide in Tanzania. Greater household wealth is associated with higher DBM; however, achieving minimum dietary diversity mitigates the negative impact of household wealth on DBM.

**How this study might affect research, practice or policy?:** This study provide evidence that supports the hypothesis that dietary diversity might be an underrated action target for addressing DBM.

Our findings encourage the implementation of double-duty approaches that simultaneously tackle different forms of malnutrition through operations such as nutrition education interventions for mothers with young children and relevant public health nutrition programs and policies in Tanzania.

## INTRODUCTION

Countries worldwide are now experiencing a fast-evolving and more complex nutrition paradigm.^1^ Instead of focusing on a single side of malnutrition, combating all forms of malnutrition is among the top priorities of the United Nations Decade of Action on Nutrition and the Sustainable Development Goals (SDGs, Target 2.2).^2,3^ Undernutrition and overweight or obesity have been historically addressed as separate challenges affecting distinct populations with contrast risk factors.^4^ However, the changing global nutrition reality is that these two distinct forms of malnutrition frequently coexist within individuals, households, and populations, with common mechanisms (e.g., economic inequalities^5^) and consequences on health.^4^ This growing recognition in the global health community forms the basis of the emerging concept of double burden of malnutrition (DBM).^6,7^ This global double burden of undernutrition and obesity and its great developmental and socioeconomic impact have been recognized as serious and lasting in low- and middle-income countries (LMICs) undergoing rapid nutrition transition^8–10;^ however, thay have not yet been examined extensively.

Tanzania is experiencing improved food availability as its economy is growing rapidly. Economic transition with an increased average household income enables more households to purchase more food^11^, which potentially improves undernutrition. However, the rates of decline in undernutrition (e.g., stunting from 34.4% in 2014 to 31.8% in 2018) in children under age five in Tanzania are still too slow to meet the SDG targets by 2030.^12^ Even worse, the prevalence of underweight increased from 13.7% in 2014 to 14.6% in 2018.^12^ Simultaneously overweight and obesity is rapidly growing, affecting over 40% of Tanzanian women aged 15–49 years,^12^ perhaps mainly due to very rapid changes in the food system in Tanzania (e.g., the availability of cheap ultra-processed fast food and beverages).^13^ This co-occurrence of undernutrition and obesity may increase DBM in Tanzania.^14^

DBM at the household-level is defined as multiple family members affected by different forms of malnutrition.^8^ Household-level DBM varies between countries and often arises in lower-middle-income countries including Tanzania^15^. Evidence showed that the prevalence of the total household-level DBM ranged between 3% and 35% across 126 LMICs, with stunted child-overweight mother pairs being the most prevalent DBM type (ranging between 1% and 24%).^15,16^ Household-level DBM has been shown to be primarily driven by socioeconomic inequalities; however, the effect of household economic status on DBM is heterogeneous.^17–21^ In poorer LMICs higher household economic levels were linked to increased odds of DBM, while in richer LMICs lower household economic levels were associated with higher odds of DBM.^22^ In Tanzania, it remains uncertain how household economic inequalities are associated with DBM. A bivariate analysis in Tanzania reported a 1.4 times higher crude likelihood of DBM among richer households; however, this study did not quantify this association while accounting for important household characteristics such as place of residence.^14^

Evidence regarding the critical factors that could help explain the link between household economic inequalities and DBM is lacking^17–22^. However, some dietary factors have been proposed to explain this link (e.g., food expenditure, changing eating habits, and the replacement of powder milk with breastfeeding).^4,5,15^ Dietary diversity, a proxy indicator for diet metrics, is hypothesized to be an underrated action target for addressing DBM^23,24^, especially among populations with diets based on starchy staples^25^ including Tanzanians. Generally, dietary diversity increases as household income increases^25^, thus improving nutrient adequacy and diet-related health outcomes^26^. Paradoxically, in emerging economies increased household income can worsen diet-related health outcomes due to poor dietary diversity^27^ and other lifestyle, social, and ecological factors^4^. Whether diversified diet can mitigate the potential adverse impact of increased household wealth on DBM in Tanzania remains unknown.

We aimed to address this gap by investigating household economic inequalities in the DBM in Tanzania, expressed as a stunted child with an overweight/obese mother, and test whether dietary diversity modifies the association between household economic status and DBM. We hypothesized that higher household wealth is associated with higher odds of DBM but that this association is attenuated by a diversified diet.

## METHODS

### Data

We obtained cross-sectional data from the 2015–2016 Tanzania Demographic and Health Survey (DSH), provided by the United States Agency for International Development.^28^ The data are from nationally representative household surveys of girls and women of productive age (15–49 years) and their children born in the five years preceding the survey, using a stratified two-stage cluster sampling method. This sampling technique allowed each household to have an equal probability of participating in the survey. In the present study, we used the dataset for children under the age of five and their mothers. This dataset provides anthropometric information for each child, as well as the characteristics of the mother and household (n = 10,233).^29^ For the analysis, we included children aged 6–23 months old (n = 3,320), who were recommended by the WHO as key targets for assessing infant and young child feeding practices using diet quality indicators such as dietary diversity.^30^ We excluded children who were not alive (n = 137), children who were not living with their mothers (n = 47), and children with height missing values (n = 55). Moreover, we excluded mothers who were pregnant (n = 207) and those with missing values of weight or height (n = 7). Our final sample consisted of 2,867 child-mother pairs (weighted sample size: n = 2,850) (see Supplemental Figure S1). Ethical approval was not required because the analysis used publicly available DHS data.

### Double burden of malnutrition

DBM can occur in different scenarios, including when a child is both stunted and overweight, when a child is wasted with an overweight mother, when a child is stunted with an overweight mother, or when a child is overweight with an underweight mother.^15^ In this analysis, we defined DBM as a child being stunted with an overweight mother in the same household, as this is the most prevalent measure of household-level DBM in LMICs.^15,22^ A child was considered stunted if their height-for-age Z-score was below minus two standard deviations (-2 SD) from the WHO Child Growth Standards median Z-score. A mother was considered overweight if her body mass index (BMI) was 25 kg/m^2^ or higher. The DBM variable was coded 1 if a child was stunted and the mother was overweight, and 0 otherwise.

### Household economic status

Household economic affluence was measured using the DHS wealth index. The DHS wealth index is a composite measure of relative household economic status, constructed using household-level information on ownership of selected assets, such as television and bicycles, materials for housing construction, and type of water access and sanitation facilities.^31^ It is one of the most useful indicators of household financial well-being in LMICs where it is difficult to obtain accurate data on household income from surveys. This is because a significant portion of the population in LMICs do not receive market level transactions and engage in significant home production.^21^ A continuous measure of relative wealth (i.e., wealth index factor score) was assessed for each household using principal component analysis.^28,29^ Based on the distribution of the wealth index factor score in the whole survey sample of the 2015-2016 Tanzania, all households were categorized into five quintiles (i.e., the wealth index quintiles).^28,29^

### Dietary diversity

Dietary diversity is a commonly used indicator of diet quality estimated using the number of different food groups consumed over a given reference period.^30^ Minimum dietary diversity was defined according to the DHS statistics guide as feeding the child with ≥ 5 out of the following eight food groups during the day or night preceding the survey: breastmilk; grains, roots, and tubers; legumes and nuts; dairy products (infant formula, milk, yogurt, cheese); flesh foods (meat, fish, poultry and liver/organ meats); eggs; fruits and vegetables rich in vitamin A; and other fruits and vegetables.^32^ The participants were categorized into two groups: achieving and not achieving minimum dietary diversity.

### Covariates

We considered the following demographic and socio-economic covariates that may affect both household economic status and the presence of DBM: the mother’s age (in years), education (no completed education, completed primary education, or completed secondary education and above), marital status (never married, currently married, formerly married), place of residence (urban or rural), whether the mother was currently breastfeeding a child (yes or no), number of children in the household, child’s age (in months) and sex (male or female), and the number of household members.

### Statistical analysis

All statistical analyses were performed using the SAS software (version 9.4; SAS Institute, Cary, North Carolina, USA) and R version 4.3.0 (R Foundation for Statistical Computing). All analyses were weighted using sampling weights, which considered the stratified cluster sampling design and non-response rate. The prevalence of DBM by region was illustrated as a choropleth map, and regional differences were tested using χ^2^ tests. We summarized the sample characteristics according to quintiles of the wealth index among non-achievers and achievers of minimum dietary diversity. Descriptive statistics were presented as weighted means and standard errors (SEs) for continuous variables, and weighted frequencies (%) and their SEs for categorical variables. We compared the sample characteristics among quintiles of household wealth index using χ^2^ tests for categorical variables and one-way analysis of variance for continuous variables.

We built logistic regression models for stratified cluster sampling to assess the odds ratios (OR) and 95% confidence intervals (CI) of DBM according to the household wealth index levels. Given that there were two few cases of DBM among child–mother pairs who achieved minimum dietary diversity in the poorest group to build logistic regression models, we combined the poorest group with the poorer group. We used both a continuous estimate of the wealth index score and groups of the wealth index as independent variables in separate models. Firstly, we performed unadjusted analyses. We then adjusted the models for covariates at baseline, namely, the mother’s age and education, marital status, place of residence, whether the mother was currently breastfeeding a child, number of children in the household, child’s age and sex, and the number of household members. We tested the trend in the association between wealth index and DBM by assigning ordinal numbers (0, 1, 2, and 3) to the groups of the wealth index categories and modeled this as a continuous variable. Based on the hypothesis that the household wealth index might exhibit varying associations with DBM depending on the presence of minimum dietary diversity, we initially tested the heterogeneity in the associations between the two groups of minimum dietary diversity. This was achieved by adding a multiplicative interaction term (minimum dietary diversity × household wealth index) to the multivariable-adjusted model. We tested this interaction effect using the likelihood ratio test by comparing the log-likelihood of the model containing the interaction term and that of the model not containing the interaction term. Statistical tests were two-sided, and a *p-*value for the interaction term < 0.05 was considered statistically significant. We conducted primary analyses separately for non-achievers and achievers of minimum dietary diversity. We performed a restricted cubic spline analysis without assuming a linear association between the wealth index score and the DBM to visualize the shape of this association. We placed four knots at the 20th, 40th, 60^th^, and 80th percentiles of the wealth index score. The 20th percentile was set as the reference value. Collinearity between independent variables was checked using the variance inflation factor test.

We performed two sensitivity analyses: (1) additionally adjusting for region to account for the potential confounding effect in which the association between household wealth and DBM is attributed to regional differences only and (2) removing the variable of place of residence from the covariates to address the potential collinearity between place of residence and the household wealth index (with a variance inflation factor value = 2.3).

## RESULTS

The estimated prevalence (SE) of DBM (stunted children with overweight/obese mother) was 5.6% (0.6) in the whole sample. The prevalence of child stunting and mother overweight was 21.3% (1.0) and 31.1% (1.2), respectively. Figure 1 shows the regional distribution of the DBM prevalence in Tanzania, which ranged from the lowest rate of 0.6% in Manyara to the highest rate of 12.2% in Kusini Unguja, with significant regional differences (*p* = 0.03). In total, 21.3% of the sample achieved minimum dietary diversity. The estimated prevalence (SE) of DBM was 5.8% (0.5) among non-achievers of minimum dietary diversity and 6.9% (1.0) among achievers of minimum dietary diversity.

**Figure 1.**
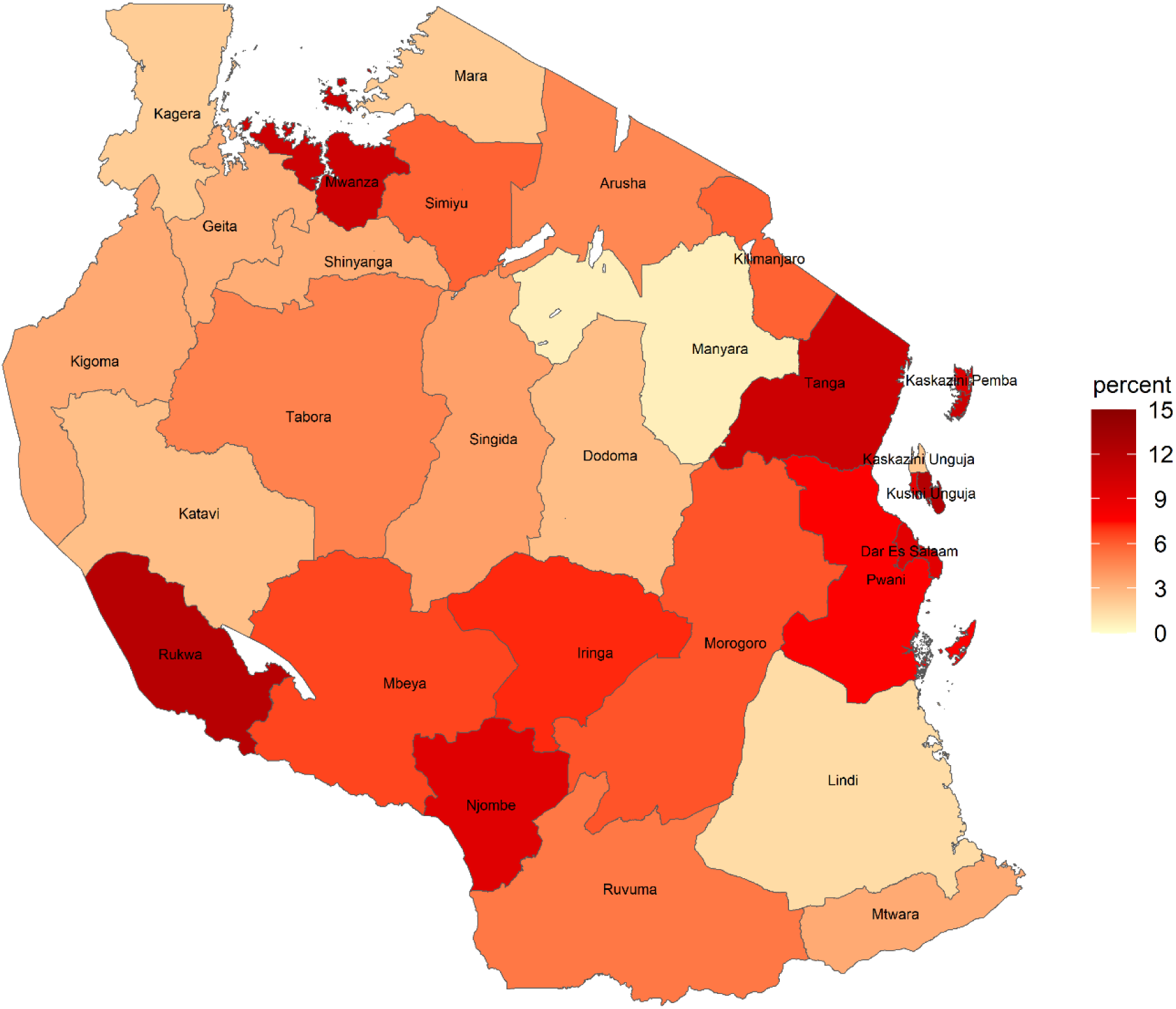
Th estimated prevalence of double burden of malnutrition (stunted child and overweight mother) in Tanzania by region

Table 1 shows the characteristics of child–mother pairs according to the quintiles of the household wealth index among non-achievers and achievers of minimum dietary diversity. In both groups, households with a higher wealth index were more likely to have mothers with higher education, had few living children and household members, lived in urban areas, and were less likely to breastfeed. They were also more likely to have children with lower mean values of height-for-age and mothers with higher mean values of BMI. In the group that did not achieve minimum dietary diversity, households with a higher wealth index were more likely to have mothers who were younger, never married, or formerly married.

**Table 1.**
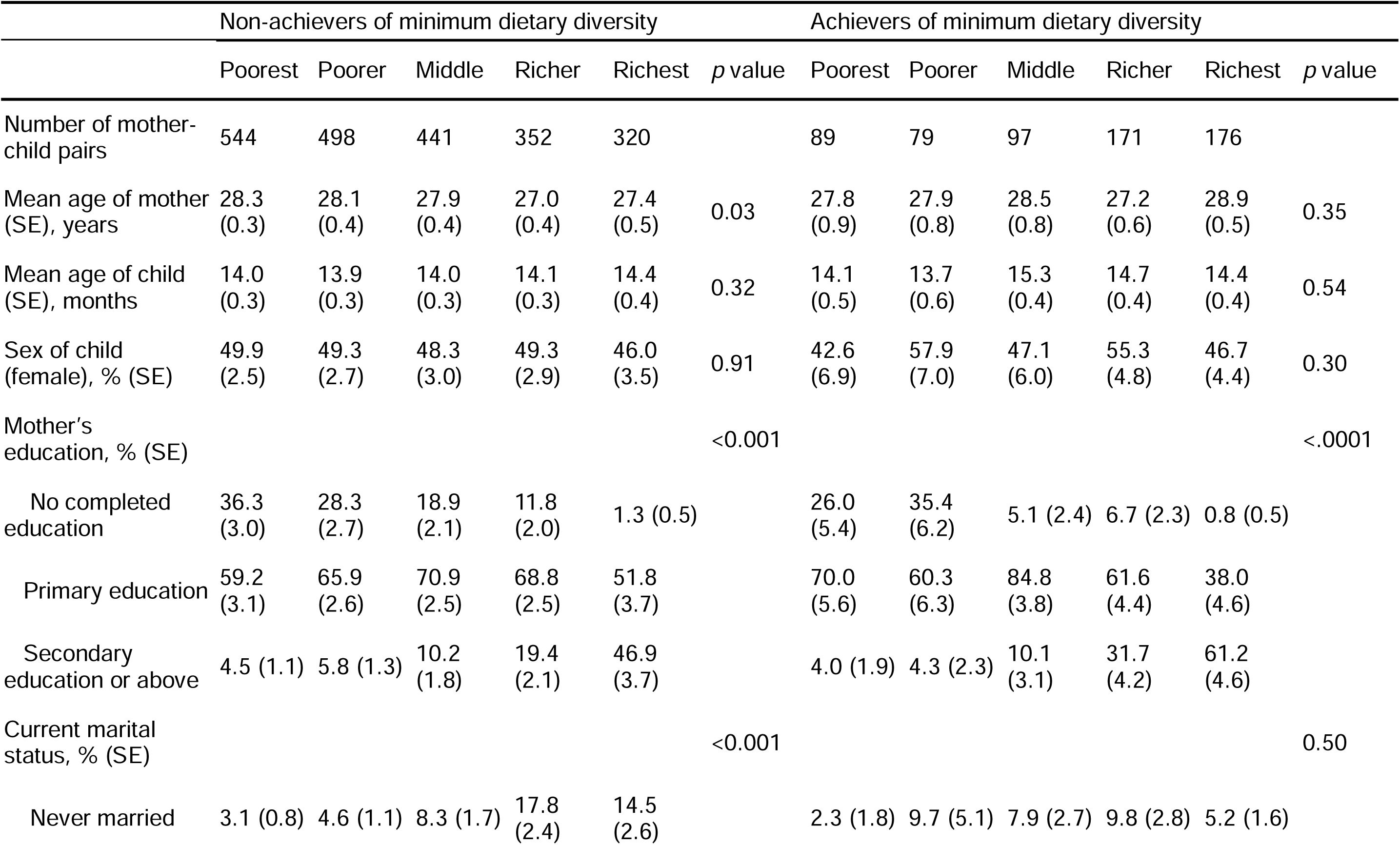
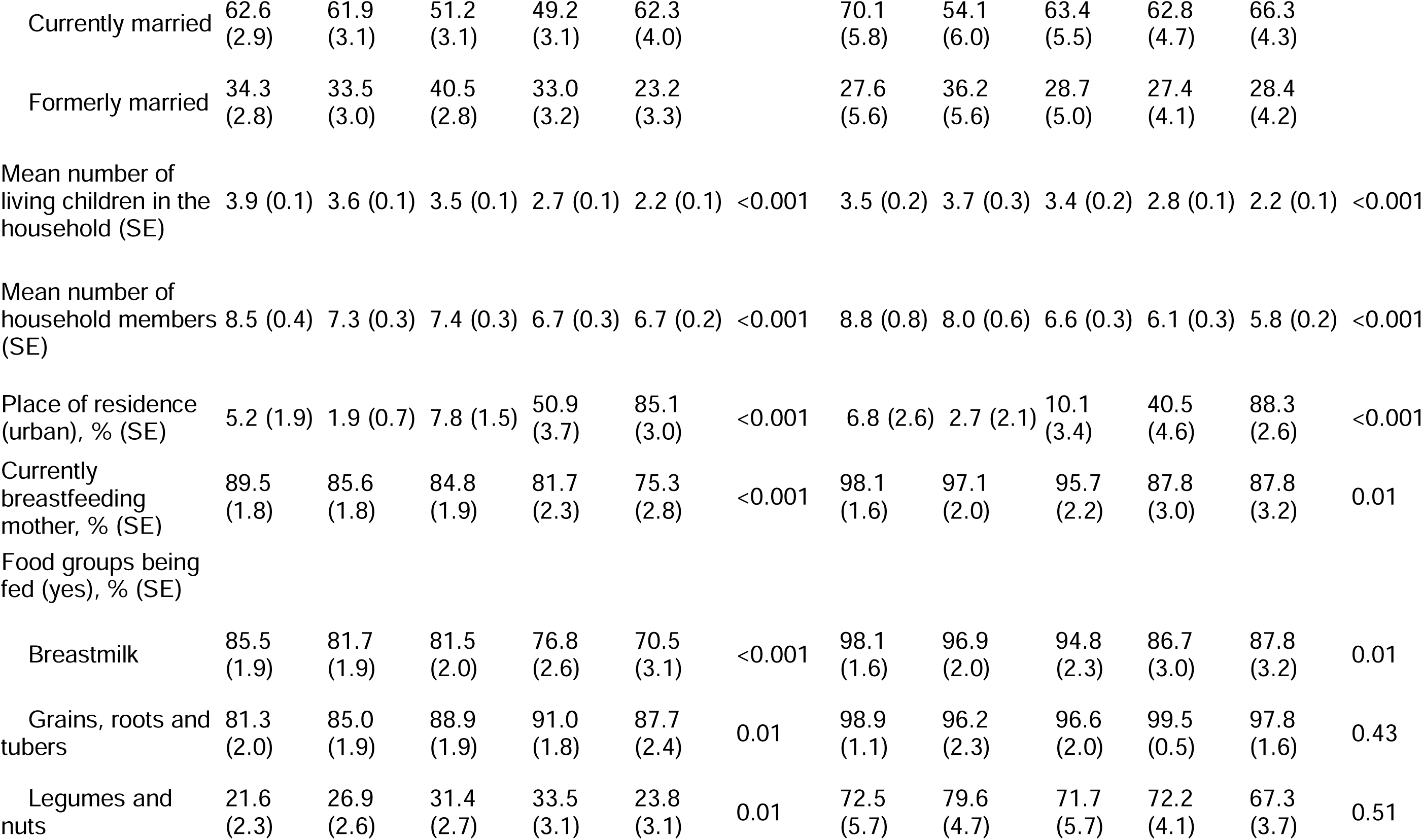
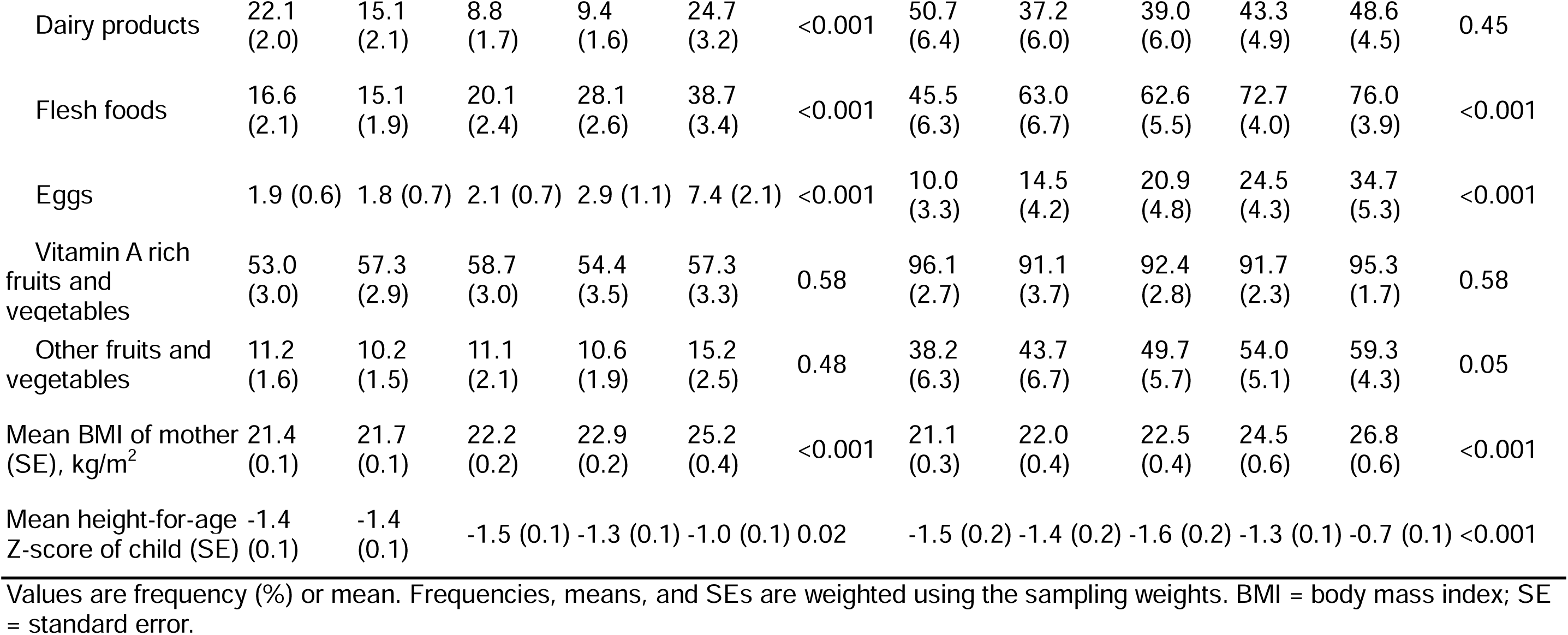
Characteristics according to the household wealth index among non-achievers and achievers of minimum dietary diversity.

Figure 2 shows the prevalence of DBM according to the household wealth level among who achieved minimum dietary diversity and those who did not. The prevalence of DBM showed a statistically significant increase with increasing household wealth index among those who did not achieve minimum dietary diversity. However, this was not observed among achievers of minimum dietary diversity. The DBM prevalence reached a plateau in the richer group and then deceased in the richest group.

**Figure 2.**
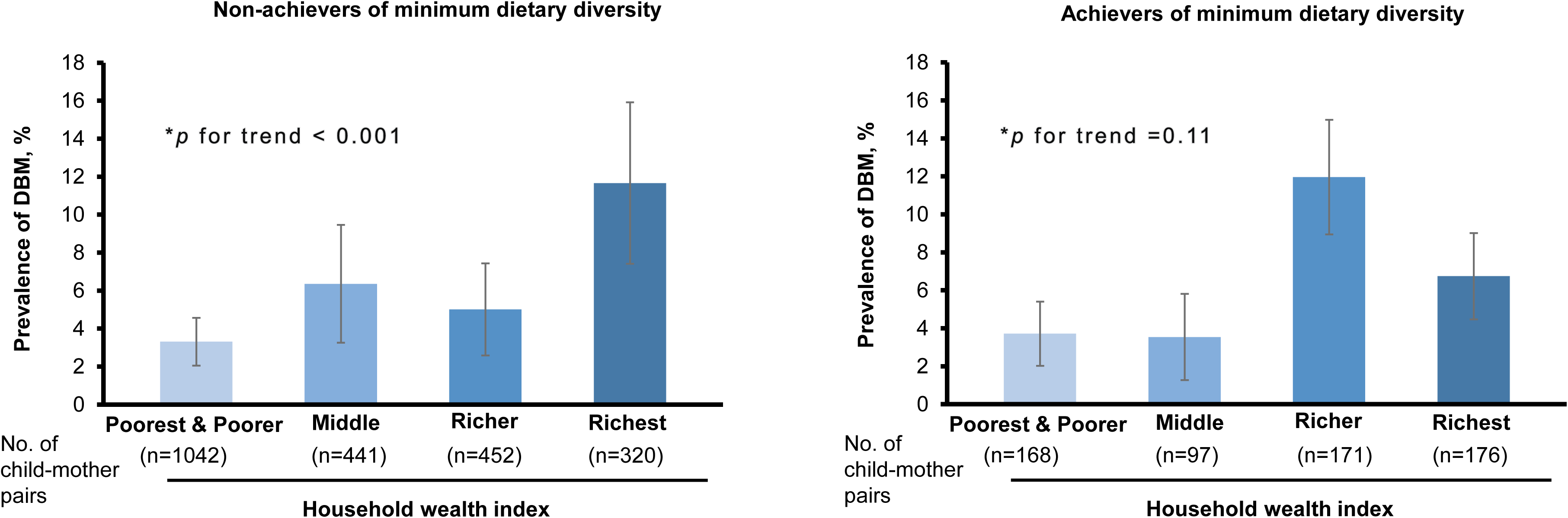
The estimated prevalence and 95% confidence interval of DBM accorind to the household wealth index levels among non-achievers and achievers of minimum dietary diversity The error bar denotes 95% confidence intervals of the prevalence. The poorest group was combined with the poorer group as there was only 1 case of DBM in the poorest group among those who achieved minimum dietary diversity. *The trend of the association was assessed by assigning ordinal numbers to each group of the household wealth index and modeling this variable as a continuous variable. DBM=double burden of malnutrition.

Table 2 shows the association between the household wealth index and DBM in unadjusted and multivariable-adjusted models. The interaction tests showed the associations of household wealth index with DBM significantly differed by minimum dietary diversity, with *p* for interaction < 0.05 in both unadjusted and adjusted analyses. Specifically, the multivariable-adjusted odds of DBM in non-achievers of minimum dietary diversity were approximately two times higher for the middle and richer groups, and more than five times higher in the richest group, as compared with the combined poorest and poorer groups (*p* for trend = 0.02). However, the multivariable-adjusted odds of DBM among achievers of minimum dietary diversity were not statistically different in the middle and the richest groups, but they were approximately five times higher in the richer group. Similar results were observed when modeling the continuous variable of the wealth index score (mean [SD]: 0.16 [0.94]) as the independent variable instead of the categories of the wealth index, with an OR (95%CI) per unit increase in the wealth index score of 2.05 (1.32-3.19) in non-achievers of minimum dietary diversity and an OR (95%CI) of 1.39 (0.76-2.54) in achievers of minimum dietary diversity. Restricted cubic spline analyses showed a similar shape association between the wealth index score and DBM among non-achievers and achievers of minimum dietary diversity (see Supplemental Figure 3). The prevalence of child stunting decreased as household wealth increased, especially among achievers of minimum dietary diversity. However, the prevalence of mother overweight significantly increased with the household wealth levels in both non-achievers and achievers of minimum dietary diversity (see Supplemental Figure S2).

**Table 2.**
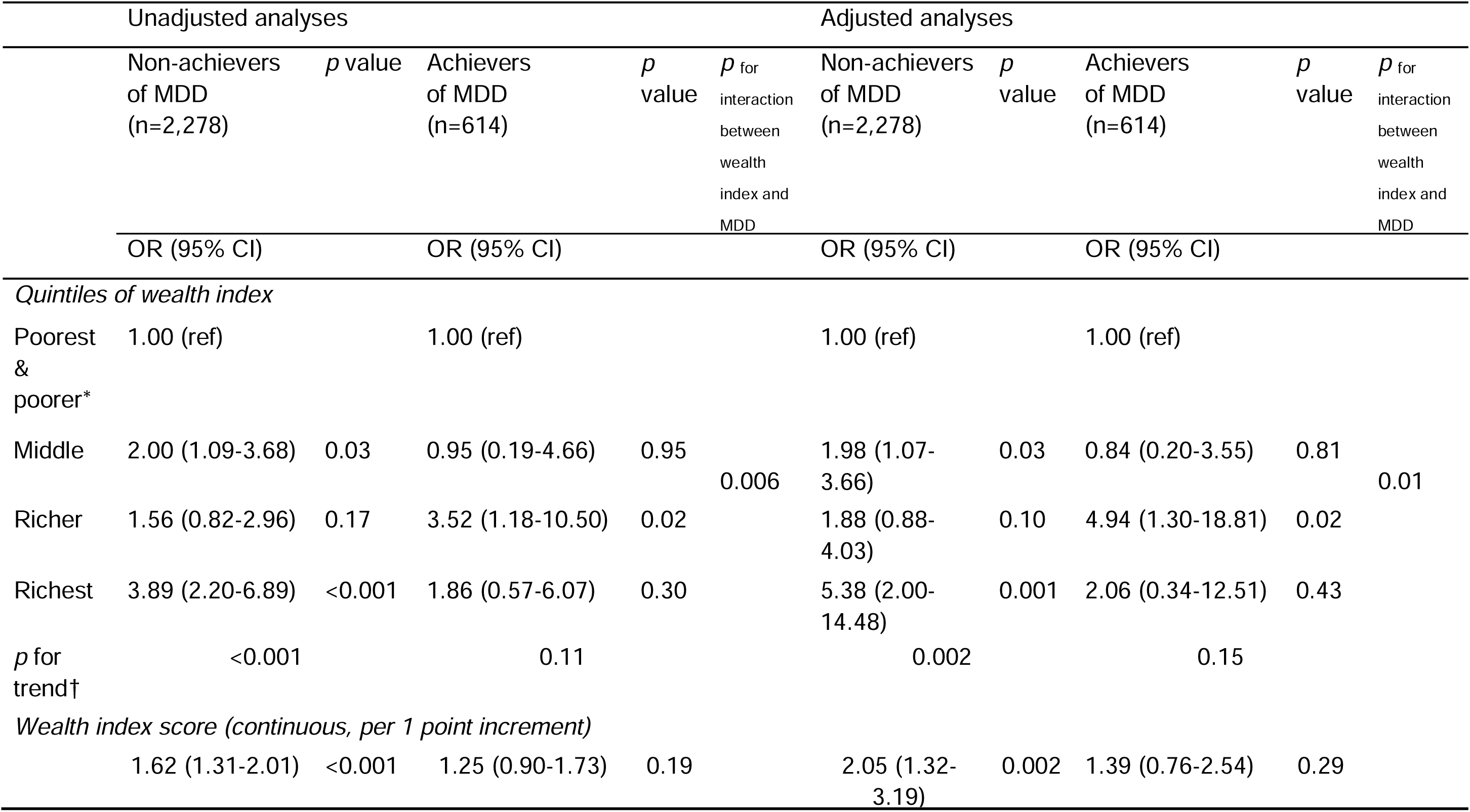
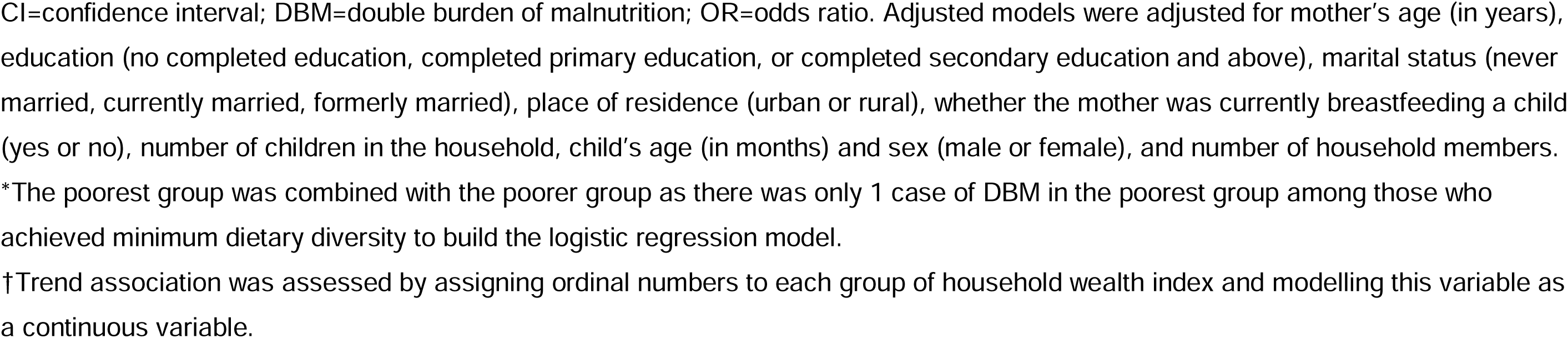
Associations between household wealth index and the double burden of malnutrition (stunted child with overweight/obese mother) among non-achievers and achievers of minimum dietary diversity (MDD)

In the sensitivity analyses, the results did not materially change after further adjustment for region (see Supplemental Table S1) or removing the variable of place of residence from the covariates (see Supplemental Table S2).

## DISCUSSION

This analysis demonstrated that the prevalence of household-level DBM varied regionally and was unequally distributed across levels of relative household wealth nationwide, and that these inequalities in DBM across levels of household wealth were moderated by minimum dietary diversity. Specifically, higher levels of relative household wealth were associated with higher odds of DBM when minimum dietary diversity was not achieved. This association was attenuated by achieving minimum dietary diversity. Our analysis quantified the association between household wealth and DBM among non-achievers and achievers of minimum dietary diversity, suggesting that household wealth increased DBM in the richest households in Tanzania; however, dietary diversity could potentially mitigate this negative impact. This study is one of the few attempts to examine the economic inequalities in DBM at the household level in Tanzania by considering the moderating role of dietary diversity in these inequalities.

Our observation of the prevalence of DBM (stunted child and overweight mother) at the household level in Tanzania is comparable to the results from analyses of LMICs (5.6% vs. 6.0%).^22^ However, our observations indicate relatively higher rates of DBM compared to LMICs in Asia, where the prevalence is mostly less than 1%.^21^ This disparity may be primarily driven by the higher prevalence of maternal overweight in Tanzania. We also observed regional differences in the prevalence of DBM; DBM tended to be disproportionately concentrated in regions with relatively high economic development levels, such as Kusini Unguja, Mwanza, Tanga, and Dar es Salaam etc. This observation supports the assumption of the undesirable impact of macroeconomic development on DBM.^21^ Our findings suggest the importance of accounting for regional variations into addressing DBM in Tanzania.

Our findings on the negative impact of household economic affluence on household-level DBM in Tanzania agree with findings from previous limited analysis of household level DBM in 55 LMICs worldwide^22^ and 11 LMICs in Asia^21^, as well as other analyses using nationally representative data^19,20^. Nevertheless, other analyses showed no or opposite direction of the association^17,18^. Of note, no previous analysis has examined the interaction between household economic affluence and dietary factors in DBM. This study expands on existing evidence regarding the adverse impact of household wealth on DBM and demonstrates that dietary diversity could potentially alleviate these negative impact. This moderating effect of dietary diversity could be driven by the observed dramatic decrease in the child stunting rate among the richest households that embraced a minimum level of dietary diversity. A more diverse diet highly is correlated with higher intake of micronutrients among children, thus helping prevent child stunting.^25^ The dramatic decrease in child stunting in the richest households could be attributed to the fact that their mothers were more likely to have a higher level of nutritional literacy, in addition to more food expenditure to sustain a high overall quality of diet for their children.^33^ We also observed a persistent increase in maternal overweight as household wealth increased, even among the group who achieved minimum dietary diversity. This result indicates that affluent Tanzanian women have a high level of total energy intake, regardless of dietary diversity. This finding could be partly explained by the cultural beliefs among Tanzanian women that associate overweight or obesity with beauty and consider it a symbol of success in life.^34^ Our findings collectively support the hypothesis that dietary diversity might be an underrated action target for addressing DBM.^23,24^

A recent Lancet Commission advocated double-duty actions to simultaneously address different forms of malnutrition, aligning with the United Nations’ SDGs and global nutrition targets.^23,24^ However, the common mechanisms of DBM through which household economic affluence affects DBM have yet to be identified. Hypothesised pathways include impact on dietary quality, food environment, physical inactivity at transportation, and breastfeeding practices.^4,13,15^ For instance, in emerging economies like Tanzania, high household income can lead to consuming fast and ultra-processed food with low-nutrient density and shifting into unhealthy eating habits^35^, thereby worsening diet-related health outcomes.^27^ Our findings indicate that double-duty actions promoting dietary diversity for children while simultaneously reducing total energy intake among mothers could be an effective strategy to address DBM in Tanzania. Additionally, the design of such double-duty actions should consider the uneven impact of economic affluence, as well as cultural and regional differences.

This study used a large nationally representative sample that provided robust estimates of the prevalence of DBM and relavent associations and enhanced the generalizability of our findings. We employed robust methodological approaches, including examining interactions between household wealth and dietary diversity and employing restricted cubic splines to avoid assuming linear associations. Our results remained robust under different sensitivity analyses, including further adjustment for region. However, this study has several limitations. The cross-sectional nature of the data precludes causal inferences. We did not include children aged two years and obove because the DHS employed the WHO-designed indicator of minimum dietary diversity specifically for children 6–23 months^30^. Future studies should validate our findings among children aged 24–59 months and their mothers in LMICs using standardized indicators.^36^ We used minimum dietary diversity for children as a proxy indicator at the household level since variables regarding the mother’s diet were not available. In Tanzania, there is a food culture in which women and children eat from the same pot ^37^, indicating that what mothers eat is strongly related to what their children eat.^38^ Nevertheless, we could not rule out the possibility of misclassification of household minimum dietary diversity in our analysis, which may have led to an underestimation of the moderating effect of dietary diversity on DBM. While our study focused on the most prevalent form of DBM, it is crucial to recognize that other forms of DBM may exhibit different associations with household economic status. The wealth index is a country-specific and relative measure of household wealth affluence. We urge caution when generalizing our findings to other countries.

In conclusion, the prevalence of household-level DBM was unequally distributed across regions of Tanzania and increased with higher household wealth. However, achieving minimum dietary diversity moderated the economic inequalities of DBM. Improved household economic status may increase DBM, but guaranteeing minimum dietary diversity could potentially address this negative impact. Our findings support and encourage the implementation of double-duty approaches that simultaneously tackle different forms of malnutrition through operations such as nutrition education interventions for mothers with young children and relevant public health nutrition programs and policies in Tanzania.

## Contributors

SC conceptualized and designed this study. SC performed the statistical analysis. SC, YS, and TH contributed to verification of the statistical results. SC, YS, TH, DLB, and BM contributed to the interpretation of the results. All authors had access to the data in the study. SC had primary responsibility for the final content. SC wrote the first draft of the manuscript, and all authors contributed to the manuscript revision.

## Funding and disclaimer

This work was supported by JSPS KAKENHI 23K16471 to SC. The funders had no role in the analysis and interpretation of data, the writing of the report, or in the decision to submit the paper for publication.

## Patient and public involvement

Patients and/or the public were not involved in the design, or conduct, or reporting, or dissemination plans of this research.

## Map disclaimer

The map included in this research paper are for illustrative purposes only. The boundaries, names, and designations used in the maps do not imply official endorsement or acceptance by the authors or affiliated institutions. The depiction of any specific geographic area, including political boundaries, does not imply any position regarding legal or political status. Users are advised to exercise caution and consult additional reliable sources for precise and up-to-date information. The authors and affiliated institutions bear no responsibility for any errors or omissions in the maps or for any consequences arising from the use or interpretation of the information presented.

## Data availability statement

This study used publicly available data that can be found online on their respective repositories: Demographic and Health Survey program (https://dhsprogram.com/). The compiled datasets, analysis files, and logs produced for this study are available from the corresponding author upon request.

